# Steroid-responsive encephalitis in Covid-19 disease

**DOI:** 10.1101/2020.04.12.20062646

**Authors:** Andrea Pilotto, Silvia Odolini, Stefano Masciocchi S, Agnese Comelli, Irene Volonghi, Stefano Gazzina, Sara Nocivelli, Alessandro Pezzini, Emanuele Focà, Arnaldo Caruso, Matilde Leonardi, Maria Pia Pasolini, Roberto Gasparotti R, Francesco Castelli F, Nicholas J. Ashton, Kaj Blennow, Henrik Zetterberg, Alessandro Padovani

## Abstract

Covid-19 infection has the potential for targeting the central nervous system and several neurological symptoms have been described in patients with severe respiratory distress. Here we described the case of a 60-year old subject with SARS-CoV-2 infection but only mild respiratory abnormalities who developed an akinetic mutism due to encephalitis. MRI was negative whereas EEG showed generalized theta slowing. CSF analyses during the acute stage were negative for SARS-CoV-2, positive for pleocytosis and hyperproteinorrachia, and showed increased IL-8 and TNF-α concentrations while other infectious or autoimmune disorders were excluded. A progressive clinical improvement along with a reduction of CSF parameters was observed after high-dose steroid treatment, thus arguing for an inflammatory-mediated brain involvement related to Covid-19.

## Introduction

Covid-19, caused by severe acute respiratory syndrome coronavirus 2 (SARS-CoV-2), is characterized by respiratory tract symptoms with possible severe outcome mostly related to pneumonia and severe acute respiratory distress syndrome (SARS)^1^. Data regarding specific neurological involvement due to SARS-CoV-2 infections are lacking. Neurological symptoms including headache and delirium have been described in epidemiological studies in up to 30% of patients^2^. Although previous coronavirus infections have been associated with involvement of the central nervous system (CNS)^3^, the neurotropism of Covid-19 is still under debate^4-6^.

The present report describes a case of encephalitis in a patient with Covid-19. During his hospitalization, he never developed SARS but had mild respiratory symptoms.

## Methods

The patient provided an informed written consent for the report and the diagnostic/therapeutic procedures were conducted in accordance with institutional and international guidelines for protection of human subjects. The study was approved by the local ethics committee of the ASST Spedali Civili di Brescia Hospital and the requirement for informed consent was waived by the Ethics Commission (NP 4051, approved 06.04.2020).

## Results

An otherwise healthy 60-year-old man presented to the Emergency Department for a severe alteration of consciousness. According to the relatives, the symptoms started five days earlier with the development of progressive irritability, confusion and asthenia followed two days later by fever, cough and cognitive fluctuations. At admission, his vital parameters were within the normal ranges and his body temperature was 36.8 °C. Oxygen saturation was within normal limits with a slight reduction of the arterial partial pressure of oxygen (Sp02 73 mmHg).

The patient showed a severe akinetic syndrome associated to mutism; he was uncooperative and unable to carry out even simple orders if not stimulated. A positive palmomental and glabella reflexes with moderate nuchal rigidity was detected with no focal signs at the neurological examination.

Blood analyses revealed normal blood cell counts, increased D-dimer (968 ng/mL) but normal levels of CRP, fibrinogen and ferritin. Chest x-ray showed moderate bilateral interstitial pneumonia. Based on the history, the radiological findings and the Covid-19 outbreak in the region, a real-time reverse transcriptase-polymerase chain reaction assays (RT-PCR) on nasopharyngeal swab was performed and confirmed a SARS-CoV-2 infection. Antiviral therapy with Lopinavir/Ritonavir 400/100 mg BID and hydroxychloroquine 200 mg BID was started.

A brain CT scan was unremarkable, and the patient was hospitalized. At admission, a lumbar puncture was performed, and CSF showed inflammatory findings with mild lymphocytic pleocytosis (18/uL) and moderate increase of CSF protein (69.6 mg/dL). The analysis of neurotropic viruses (HSV-1, HSV-2, HSV-6, HSV-8, EBV, VZV, Adenovirus and Enterovirus) was negative. CSF RT-PCR for SARS-CoV-2 was also negative. An electroencephalography (EEG) exhibited generalized slowing with decreased reactivity to acoustic stimuli (Figure 1). The same day an empirical treatment of ampicillin and acyclovir was introduced. The day after, the magnetic resonance imaging (MRI) with gadolinium contrast did not reveal any significant alterations or contrast-enhanced areas within the brain and/or meninges (Figure 1). Three days after admission, given the persistence of clinical symptoms, high-dose intravenous steroid treatment was started (methylprednisolone 1 g/day for five days). CSF analyses, carried out one day after steroid administration, showed lymphocytic pleocytosis (18/uL), hyperproteinorrachia (127.2 mg/dL) and normal Link index without oligoclonal bands. CSF RT-PCR for SARS-CoV-2 was still negative. At this time, CSF analyses revealed relatively slightly increased levels of interleukin 6 (IL-6) of 2.36 pg/mL, strongly increased levels of interleukin 8 (IL-8, higher than 1100 pg/mL) and increased levels of tumor necrosis factor alpha (TNF-α, 1.31 pg/mL) and β2-microglobulin (β2M, 3.06 mg/L). Biomarkers for neuronal injury (tau and neurofilament light [NfL]) were normal (Supplementary Table 1). After the first infusion, the patient improved-He became alert and able to execute simple tasks on command and to repeat single words. The following days, the patient was able to talk and to answer simple questions. Once the five days-high dose steroid therapy ended, he displayed only mild disinhibition and somewhat fluctuating alertness.

**Figure 1.**
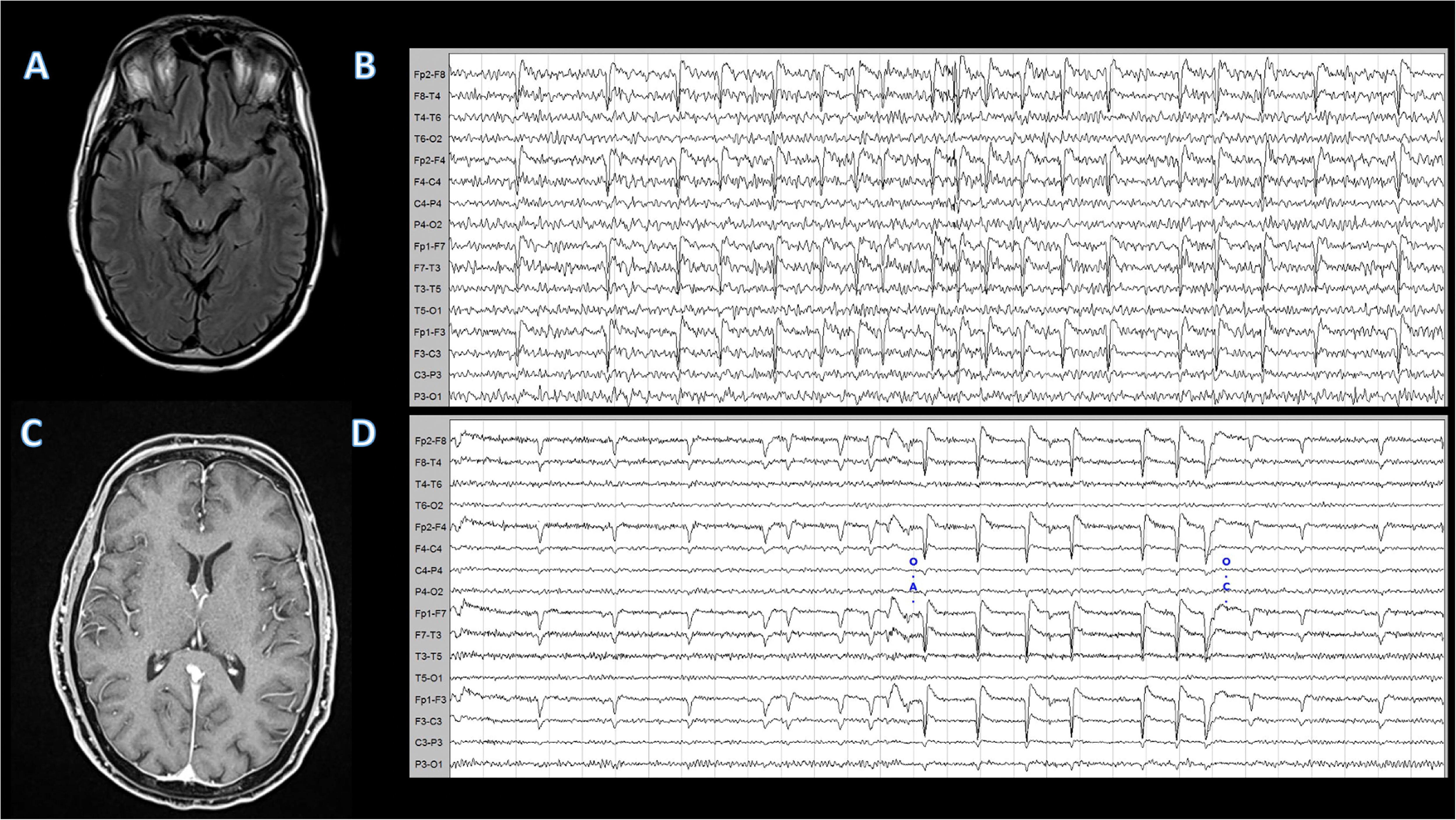
Magnetic resonance imaging (MRI) and electroencephalography (EEG) findings. Box A/C normal MRI findings on T2 (A) and T1 with gadolinium (C). EEG alterations at admission during the akinetic mutism state with global mid-amplitude theta slowing without reactivity to eye opening (B) and reorganization of the background alpha rhythm and reactivity after 5 days of high-dose steroid treatment (D). Electroencephalographic acquisition settings: 10-20 system, longitudinal montage. Recording speed: 30 sec/page; Sensitivity: 7 μV/mm; time constant: 0.1 sec, high-frequency filter: 15 Hz.

**Figure 2.**
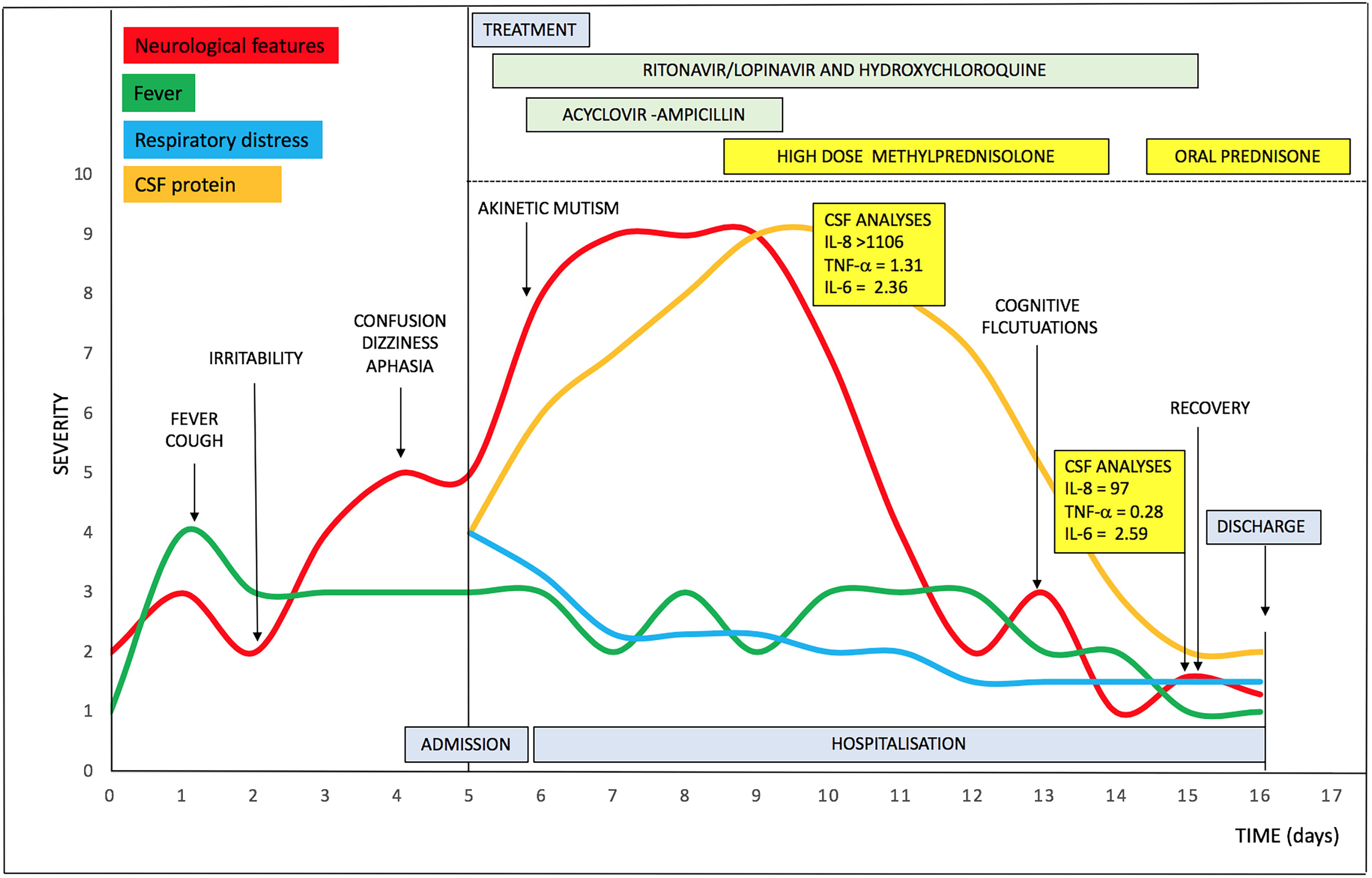
Clinical features, treatment and respiratory/inflammatory profile during the disease course. Abbreviations: CSF, cerebrospinal fluid; interleukin 6, IL-6; interleukin 8, IL-8; tumor necrosis factor alpha, TNF-α.

A follow-up MRI (9 days post-admission) was normal while serial EEG revealed improvement and normalization of reactivity and slowing (Figure 1, D). A wide immunological screening of immune-mediated encephalitis (antibodies against NMDAR, LGI1, CASPR2, GABA_A_R, GABA_B_R and AMPAR, Ri, Yo, Ma2, Hu and amphiphysin) was negative, as well as anti-thyroglobulin, anti-thyroid peroxidase and anti-MOG antibodies (Supplementary details).

Ten days after the admission, CSF protein concentrations decreased (91.4 mg/dL), but lymphocytic pleocytosis (38 cells/uL) increased. CSF IL-8 and TNF-α decreased to 97 pg/mL and 0.28 pg/mL, respectively, whereas IL-6 and β2-microglobulin values were stable (Supplementary table 1). CSF tau and NfL concentrations remained normal. Given the remaining neuroinflammatory changes, oral prednisone was started with progressive tapering. At discharge, eleven days after admission, the patients presented with normal neurological examination.

## Discussion

The potential neurotropism of SARS-CoV-2 with direct or autoimmune-mediated response is still debated^4-6,7^. We here described a case of steroid-responsive encephalopathy in a patient with confirmed SARS-CoV-2 infection.

After the first reports describing severe pneumonia cases in Wuhan, China, Covid-19 rapidly spread worldwide with critical challenges for the health care systems globally^8,9^. The World Health Organization has thus declared Covid-19 a public health emergency of international concern, with more than 200,000 deaths on the 25^th^ of April. Brescia, situated in the Lombardy region, northern Italy, was one of the provinces most affected by Covid-19 accounting for more than 2100 deaths on the 25^th^ of April 2020.

Epidemiological studies showed that Covid-19 presents in most cases with fever and upper respiratory symptoms^2^. Recently, the study performed by Mao and coauthors^6^ reported neurological manifestations of Covid-19 in the outbreak in China in up to 36.4% of patients hospitalized, including alteration of consciousness, headache, dizziness and delirium^6^. Neurological manifestations were also reported in previous epidemiological studies on larger samples^2^. Neuro-infectivity has been described for other coronaviruses but is still questioned for SARS-CoV-2. Support for a hypothesis of CNS infection through a nasopharyngeal route is provided by clinical observations of frequent and persistent anosmia/dysgeusia.

Several authors reported neurological symptoms in severe cases, supporting the concept that CNS symptoms might be secondary to severe respiratory failure^7^.

Another important aspect is the hyperinflammation state secondary to SARS-CoV-2 infection, with massive release of cytokines and chemokines that could alter the permeability of blood-brain barrier (BBB). This phenomenon could indeed cause the activation of the neuroinflammation cascade^5^. Hypothetically, SARS-CoV-2 could also induce, by molecular mimicry-related mechanisms, the production of antibodies against neural or glial cells, as demonstrated for HSV-1, EBV or Japanese encephalitis^10^.

The here described Covid-19 case is of particular interest, as the patient presented with encephalitis with only mild respiratory alterations. Meningeal signs at presentation suggested meningoencephalitis and the patient started combined antibiotic and antiviral treatment, according to current guidelines for CNS infections^11^. Lack of presence of Covid-19 in two different CSF samples could not definitely exclude CNS infection, as the presence of the virus in CSF could be transient or not detectable, such as in the cases of 40% of West Nile and up to 69% of enterovirus infections^12^. In our case, the blood brain barrier alterations supported by CSF analyses argue for a potential CNS invasion via blood route of COVID-19^3^. Of interest, peculiar akinetic mutism presentation has been associated with cases of Epstein-Barr encephalitis^13^. However, the normal MRI findings, with no meningeal enhancement or brain alterations, and the response to steroid treatment support other alternative pathophysiological mechanisms. In this case, we excluded the most common causes of autoimmune encephalitis, akinetic mutism and psychosis so far^14-16^. Another possible explanation of transitory akinetic mutism would be an abnormal neuroinflammation response induced by SARS-CoV-2^17,18^, better fitting with the prompt response to steroid treatment.

To support this hypothesis, we measured CSF levels of IL-6, IL-8, TNF-α and β2-microglobulin at two different time points. At the time of the akinetic mutism presentation, we observed pleocytosis and a severe increase of CSF inflammatory proteins, IL-8 in particular, but also TNF-α and β2-microglobulin. After clinical improvement, the inflammatory biomarkers decreased, whilst β2-microglobulin and CSF cell counts remained abnormal. This suggest a possible cytokine-mediated hyperinflammation response. The role of steroid treatment in the resolution of symptoms of the here-presented case, however, definitely need further observations to be confirmed. Indeed, a still unpublished work posted on pre-print described two cases of suspected encephalitis associated with Covid-19, in which the improvement was in a similar time frame treatment-independent^19^. Of interest, CSF cytokine alterations were not associated with severe abnormalities of peripheral markers of inflammation or severe respiratory involvement^19^, arguing for a CNS-specific abnormal inflammation response leading to neurological manifestation^20^. This fit with normal serial imaging and the absence of biomarker evidence of neuronal injury, suggesting a functional rather than a destructive neuronal network impairment.

## Conclusions

The case highlighted that Covid-19 may involve severe neurological alterations independently from respiratory function. Despite the lack of a clear pathophysiological mechanism, the apparently positive response to steroid treatment together with the normalization of CSF cytokines argues for the encephalitis in our patient to have been mediated by a hyperinflammatory mechanism. On the other hand, although there was no evidence of SARS-CoV-2 in the CSF by RT-PCR, a direct CNS infection cannot be excluded. This report suggests that it may be useful to investigate the therapeutic effects of corticosteroid administration in Covid-19 related encephalitis.

## Data Availability

Data are available upon responsible request by contacting the corresponding author

## Acknowledgement

We thank the patient for his participation in the study.

## Author contribution

AP, SO, SM and AP contributed to the conception and design of the study, AP, SO, SM, AC, IV, SG, SN, AP, EF, AC, ML, MPP, RG, FC, NA, KB, HZ, AP contributed to the acquisition and analyses of data; AP, SO, SM, SG and AP contributed to drafting the text and preparing the figures

